# Comparative Analysis of RT-PCR, RT-LAMP, and Antigen Testing Strategies for Effective COVID-19 Outbreak Control: A Modeling Study

**DOI:** 10.1101/2024.07.14.24310377

**Authors:** Tanakorn Chantanasaro, Chayanin Sararat, Noppamas Yolai, Pikkanet Suttirat, Kawin Nawattanapaiboon, Somchai Chauvatcharin, Sudarat Chadsuthi, Charin Modchang

**Affiliations:** Biophysics Group, Department of Physics, Faculty of Science, Mahidol University, Bangkok 10400, Thailand; Center for Disease Modeling, Faculty of Science, Mahidol University, Bangkok, 10400, Thailand; Zenostic Co., Ltd, Bangkok 10400, Thailand; School of Materials Science and Innovation, Faculty of Science, Mahidol University, Bangkok 10400, Thailand; Department of Biotechnology, Faculty of Science, Mahidol University, Bangkok 10400, Thailand; Department of Physics, Faculty of Science, Naresuan University, Phitsanulok 65000, Thailand; Centre of Excellence in Mathematics, Ministry of Higher Education, Science, Research and Innovation, Bangkok 10400, Thailand; Thailand Center of Excellence in Physics, Ministry of Higher Education, Science, Research and Innovation, 328 Si Ayutthaya Road, Bangkok 10400, Thailand

## Abstract

The COVID-19 pandemic has highlighted the crucial role of testing in mitigating disease transmission. This study comprehensively evaluates the effectiveness and cost-efficiency of various testing strategies, including daily screening, symptom-based testing, and contact-based testing, using assays such as RT-PCR, RT-LAMP, and antigen tests. Employing stochastic modeling on a contact network, we assessed the impact of these strategies on outbreak control, using COVID-19 as a case study. Our findings demonstrate that daily screening, particularly with RT-PCR and RT-LAMP, significantly reduces transmission risks but incurs higher costs. In contrast, symptom-based testing offers a more cost-effective alternative, albeit with lower efficacy in mitigating outbreaks. Notably, testing turnaround time emerges as a more critical factor than assay sensitivity in containing outbreaks. Moreover, combining symptom-based testing with contact tracing further reduces outbreak probability and scale. To provide a comprehensive analysis, we also explored the application of these strategies in scenarios where a portion of the population has acquired immunity. Our results suggest that testing all symptomatic individuals is the most effective and cost-efficient approach in the later stages of an epidemic. These findings provide valuable insights for optimizing testing strategies to tackle current and future infectious disease outbreaks effectively and efficiently. By adapting strategies based on the stage of the epidemic, population immunity, and available resources, public health authorities can design targeted interventions to protect communities while managing limited resources.

## Introduction

The Coronavirus disease 2019 (COVID-19) pandemic has had a devastating global impact, with over 775 million cases and approximately 7 million deaths reported worldwide as of May 2024 ^1^. The pandemic has triggered a multifaceted crisis, adversely affecting the economy, tourism, and public health and exacerbating poverty ^2–8^. In response to this unprecedented challenge, many countries implemented non-pharmaceutical interventions (NPIs) such as physical distancing, remote work, face mask-wearing, and school and workplace closures while awaiting the widespread distribution of effective vaccines ^9–13^. However, despite the availability of vaccines, the emergence of new variants and waning immunity has led to ongoing breakthrough infections and reinfections, contributing to a persistently challenging situation ^14–18^. Consequently, NPIs remain crucial in limiting transmission and mitigating the impact of the pandemic.

Among the various NPIs, testing and isolating infected individuals has played a critical role in mitigating the burden of the COVID-19 outbreak. The rapid identification of infections through testing enables the prompt isolation of infectious individuals, thereby limiting further transmission from primary cases and reducing the overall incidence of the virus ^12,19,20^. Moreover, screening tests aimed at detecting asymptomatic or pre-symptomatic infectious individuals within the population can significantly help curtail onward transmission ^20–22^. By identifying and isolating infected individuals who may not exhibit symptoms, screening tests can break the chain of transmission and prevent the silent spread of the virus in the community.

Several approaches are employed in viral testing to detect the presence of SARS-CoV-2 ^23–25^. These approaches involve identifying specific viral components, such as the ribonucleic acid (RNA) or antigens, within an individual’s body to determine current or recent infection with SARS-CoV-2. Nucleic acid amplification testing (NAAT), which includes the widely used reverse transcription polymerase chain reaction (RT-PCR), is one such approach that detects viral RNA genes ^23,24^. In fact, RT-PCR is considered the gold standard for COVID-19 diagnosis due to its high sensitivity and specificity, making it an essential tool for accurate case detection and outbreak management. However, RT-PCR tests often require specialized equipment and trained personnel, which can limit their availability and increase costs. In contrast, antigen tests target specific viral proteins known as antigens and are often used for rapid, point-of-care testing ^23,24^. Although antigen tests may have lower sensitivity compared to RT-PCR, they offer several advantages, such as cost-effectiveness, faster turnaround times, and ease of use, making them suitable for large-scale screening and rapid outbreak control ^23,24,26^. Another promising testing method is reverse-transcription loop-mediated isothermal amplification (RT-LAMP), which is a reliable and rapid screening test that can be used in the field or under non-laboratory conditions ^27^. RT-LAMP has been shown to have high sensitivity and specificity, comparable to RT-PCR while being more cost-effective and easier to implement in resource-limited settings ^25,27^.

Modeling studies have suggested that effective testing strategies, when combined with other interventions such as contact tracing, isolation, and quarantine, have the potential to prevent both the initial epidemic and its resurgence ^20,28–31^. However, limited research has directly compared the impact of different testing strategies on outbreak risk and epidemic size ^32–34^. As such, there is a need for more comprehensive studies that evaluate the effectiveness and cost-efficiency of different testing methods and strategies across diverse settings, taking into account factors such as test sensitivity, turnaround time, and the interplay with other control measures.

In this study, we employ an individual-based modeling approach to investigate the impact of various testing strategies on mitigating COVID-19 transmission. We compare the effectiveness of daily screening, symptom-based testing, and contact-based testing in reducing outbreak risk and epidemic size. Furthermore, we examine the influence of factors such as turnaround time and assay sensitivity on the efficacy of these testing strategies. To provide a comprehensive understanding of the trade-offs involved, we also conduct a comparative cost analysis for each testing strategy in terms of outbreak control. By evaluating the interplay between testing strategies, their associated costs, and their effectiveness in curbing disease spread, this study aims to offer valuable insights for policymakers and public health authorities in designing optimal testing approaches to combat the ongoing COVID-19 transmission and future infectious disease outbreaks.

## Materials and Methods

### Viral Dynamics within Host Body

We estimated the viral load (VL) within the host body over the course of infection by converting the cycle threshold (Ct) values obtained from Chia *et al.* ^35^ using the following formula ^36^:

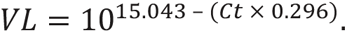

However, the Ct values prior to diagnosis were not available in the dataset. Therefore, we estimated the viral dynamics before diagnosis for both unvaccinated and vaccinated individuals by fitting the number of viral copies post-diagnosis to the innate immune response model outlined in ^37^ (**Fig. 1A**). The fitting process was conducted using a nonlinear least-squares function in MATLAB.

**Figure 1.**
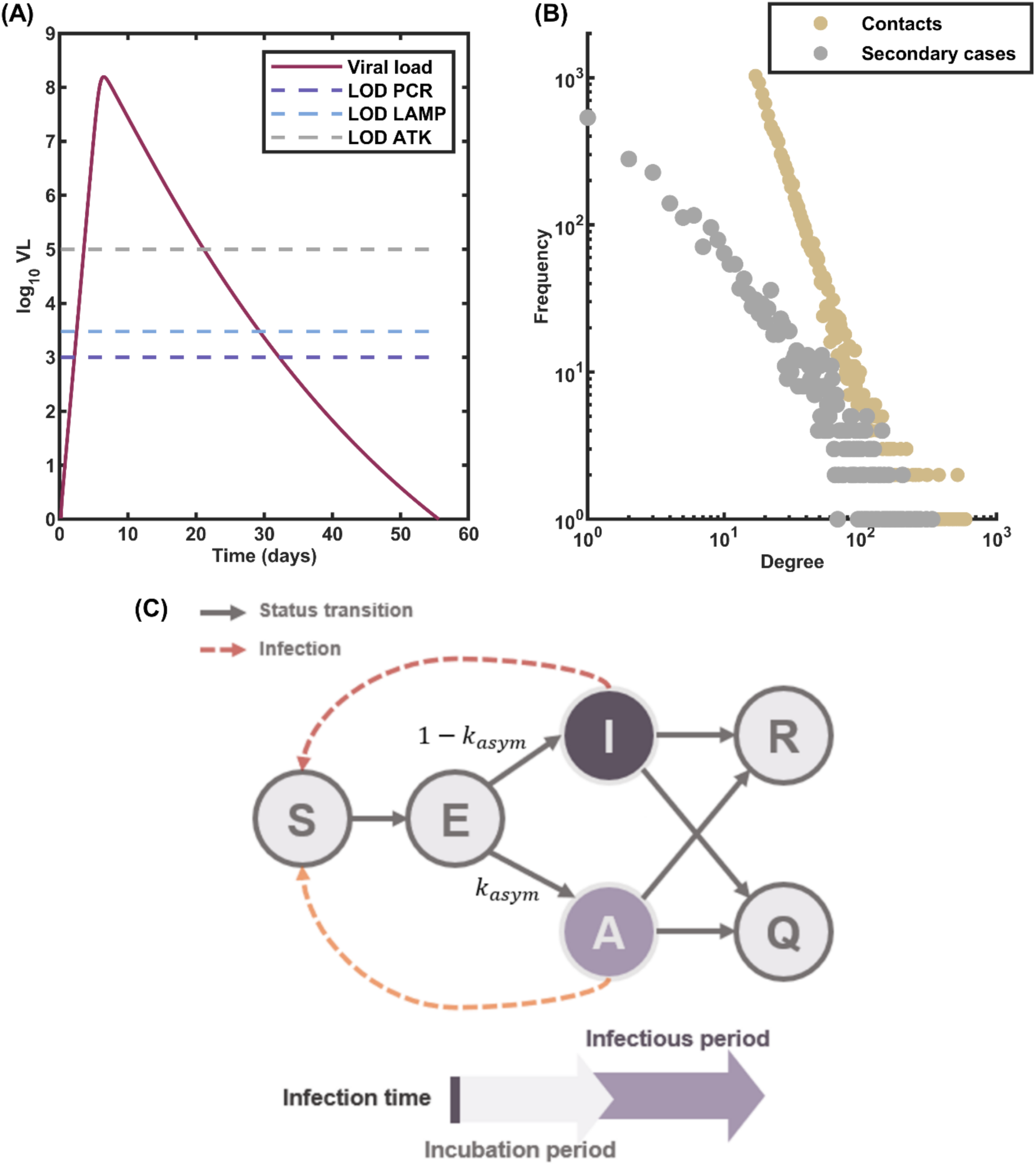
Model Structure. (**A**) Estimated viral load dynamics of an infectious individual following infection, plotted alongside the limits of detection (LOD) of three testing assays: RT-PCR, RT-LAMP, and ATK. (**B**) Distribution of the number of contacts in the simulated contact network (yellow dots) and the expected number of secondary cases resulting from an infected individual (gray dots). The contact network is generated using a power-law distribution, capturing the heterogeneity in social interactions. (**C**) Compartmental model representing the progression of an infected individual’s infection status. Solid arrows depict the transitions between different stages of infection. Dashed arrows indicate the potential for disease transmission from infectious individuals to susceptible ones.

### Infectiousness Profile

We assessed the level of infectivity exhibited by an individual throughout their infection by creating an infectiousness profile (𝑝(𝑡)). This profile was constructed using viral load (VL) values, following the methodology detailed in ^37^, and can be described as follows:

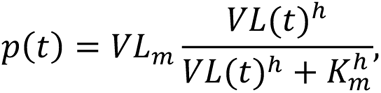

where 𝑉𝐿_𝑚_represents the maximum viral load level, while 𝑉𝐿(𝑡) is the estimated viral load as determined in the previous subsection. The constant ℎ and 𝐾_𝑚_are fixed at specific values, with ℎ set to 0.51 and 𝐾_𝑚_ set to 8.9 × 10^6^ RNA copies/mL, respectively.

### Contact Network

This study simulated the transmission dynamics within a contact network characterized by nodes exhibiting a power-law distribution in their degree. Power-law distributions are pervasive across various intricate systems, such as wealth distribution, city sizes, social networks, word frequencies, and citations ^38–42^. Studies have indicated that social interaction networks often adhere to a power-law distribution ^43,44^. Utilizing the Barabási–Albert (BA) algorithm, we constructed the network and subsequently conducted simulations to model the transmission process within it ^45^. Nodes and links within the network represent individuals and their respective contacts. The contact network comprises 10,000 nodes, with an average degree of 17 (**Fig. 1B**). This average degree choice ensures that the number of secondary cases derived from a negative binomial distribution for all individuals remains within the bounds of their actual contacts. The degree distribution of the generated network follows a power-law distribution with the power-law exponent of 2.881.

### Model Structure

In the model, each individual can possess one of six states: susceptible (𝑆), latently infected (𝐿) , symptomatic infectious (𝐼_𝑆_) , asymptomatic infectious (𝐼_𝐴_) , isolated (𝑄), and recovered (𝑅), as depicted in **Fig. 1C**. When susceptible individuals become infected, they transition to the latent state and progress through the course of infection. Once in the latent state, there is a probability, denoted as 𝑘_𝑎𝑠𝑦𝑚_, that they become asymptomatic. The asymptomatic infectious individual has a lower viral load and thus is less infectious than the symptomatic one ^46^. A portion of individuals undergo testing to determine their infection status. If a person tests positive for the disease, they are isolated from the general population until they recover.

In the transmission network, the number of secondary infections resulting from a single primary case was modeled using the negative binomial distribution with a mean equal to the basic reproduction number (𝑅_0_) and a dispersion parameter (𝑘). For the asymptomatic infectious individual, the number of secondary infections was adjusted by a constant 𝑟, which reflects the reduced infectiousness ^47^. The model parameters and their default values are summarized in **Table 1**.

**Table 1.**
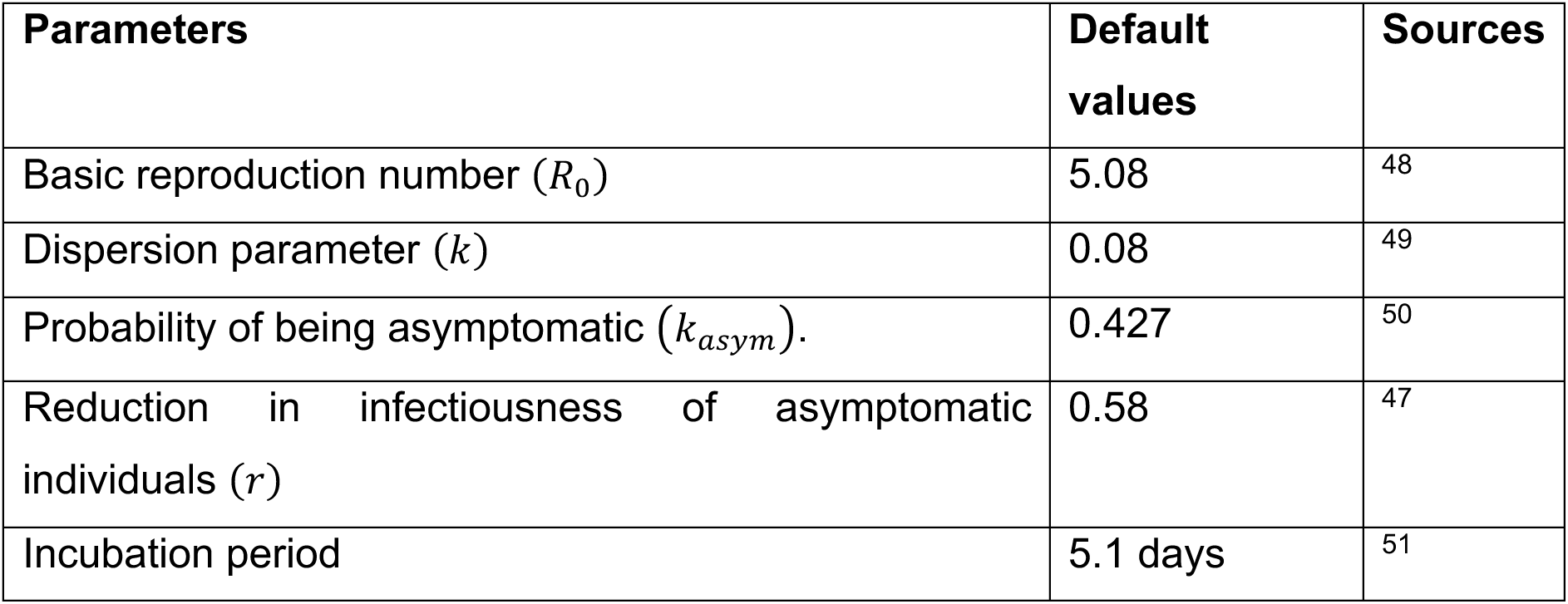
Model parameters and their default values.

### Testing Strategies

We investigated three testing strategies used to monitor and control the spread of COVID-19:

1) **Daily screening**: In this testing scenario, a fixed group of individuals is tested for infection every day. This strategy focuses on regular testing of a predetermined population to detect and isolate cases early, even before symptoms develop. As a result, asymptomatic and pre-symptomatic infectious individuals could be identified a few days after the infection.
2) **Symptom-based testing**: Symptom-based testing targets individuals who have developed symptoms of the disease, typically around day 5.1 post-infection. Since this strategy relies on the identification of symptoms as a trigger for testing, it cannot detect asymptomatic or pre-symptomatic carriers.
3) **Contact-based testing**. Contact-based testing involves testing individuals who have been in close contact with a confirmed positive case in the contact network. If any individuals in the contact network get a positive test result, their contacts within the network will be tested for seven consecutive days since the primary case is confirmed. The seven-day testing period covers the pre-transmission of the secondary cases. Note that the contact-based testing strategy alone cannot detect the initially infected individuals; therefore, symptom-based testing with a 75% chance was used in conjunction when the simulations started.

### Testing Assays

In our study, we considered the use of different testing assays, including real-time polymerase chain reaction (RT-PCR), reverse transcription-loop-mediated isothermal amplification (RT-LAMP), and antigen test kit (ATK). Each of these assays possesses distinct characteristics, making them well-suited for different applications within the testing strategies. Key differences among these assays are the turnaround time, the time delayed since the testing and the result, the limit of detection (LOD), which is the minimum viral load that the assay can detect, and their accuracy. Our modeling framework focused primarily on the differences in turnaround time and LOD. When the viral load level exceeds the LOD, we assumed a 100% probability of a positive test result. The specific values of turnaround time, limit of detection, and cost per test for each assay are listed in **Table 2**. Although RT-LAMP and ATK are known to provide results in the order of minutes in practice, we conservatively set their turnaround times to zero for modeling simplicity.

**Table 2.**
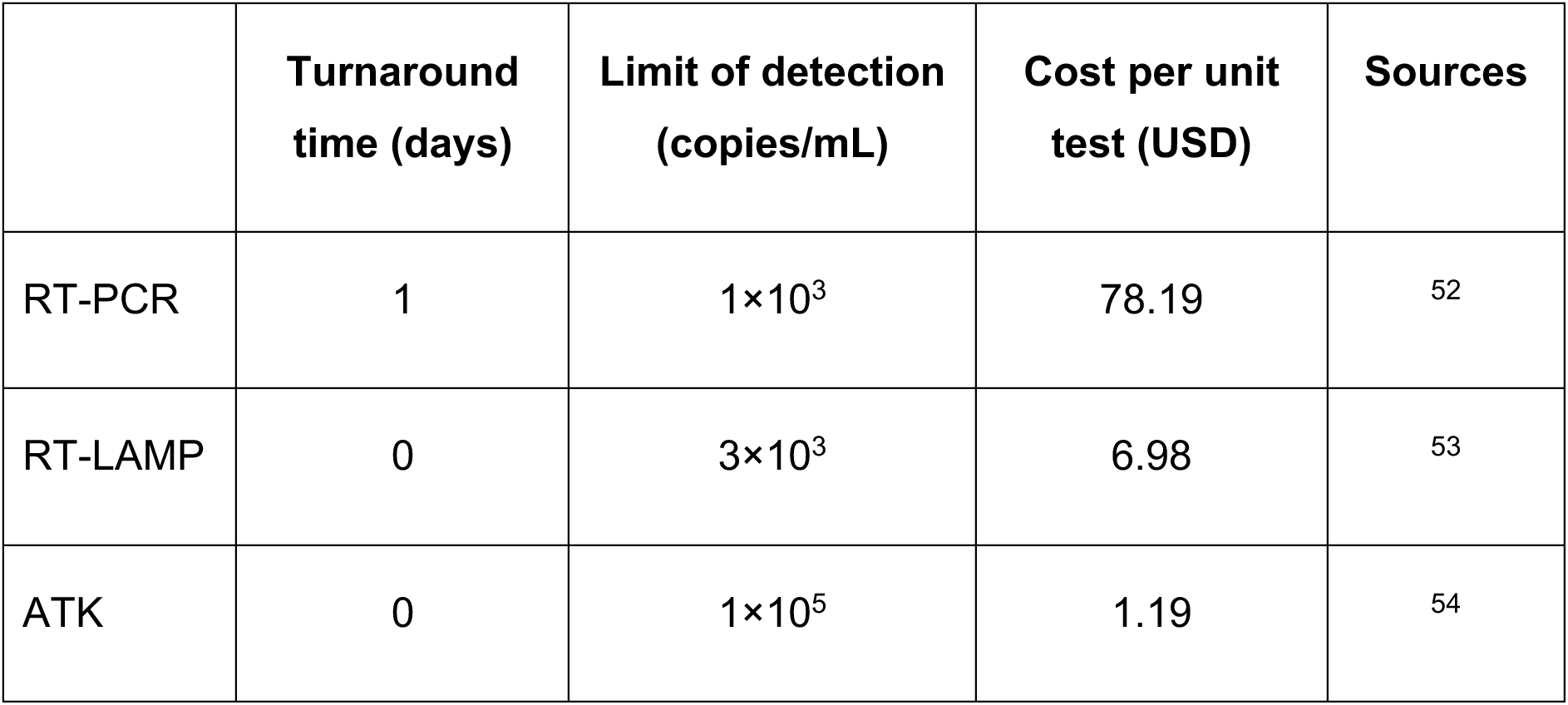
Turnaround time, limit of detection (LOD), and cost per test of each testing assay.

### Estimating the Probability of an Outbreak

The probability of an outbreak serves as a key metric for evaluating the effectiveness of various testing strategies. Effective testing strategies should minimize the risk of a widespread outbreak. To quantify the risk, we established a threshold value of cumulative cases at 1,000 at the end of the 300-day simulation. This threshold value serves as a critical point of distinction between two scenarios. One is the scenario in which the primary infectious individual spreads the disease to only a limited number of cases, and the outbreak eventually goes extinct, and the other is where the disease spreads extensively to a substantial part of the community. The probability of an outbreak is then calculated as the ratio of simulation runs that the disease spreads to a substantial part of the community to the total number of runs conducted ^55^.

### Estimating the Cost Associated with Each Testing Strategy

To estimate the cost of testing, we focused on the number of individuals isolated as a key factor. The cost calculation varies based on the testing strategy employed. For the daily screening strategy, the cost of testing for each simulation was iteratively calculated, starting with an initial cost and increasing based on the number of individuals tested and the number of new cases detected in each simulation. The testing cost is calculated until the end of the outbreak when no infected individuals are left. The cost calculation is more straightforward for the symptom-based testing and contact-based testing strategies. It is determined by multiplying the number of tested individuals by the cost of the assay per test. We then compute the average cost of testing across all simulation runs, focusing on those where at least one infectious individual was tested.

## Results

### Impact of Daily Screening on Outbreak Dynamics

To evaluate the impact of daily screening on COVID-19 transmission, we conducted simulations within a network of 10,000 individuals. Initially, we randomly selected one person to be in the latent state. Our simulations revealed that as the percentage of the tested population increased, there was a decrease in the probability of an outbreak, and the cumulative number of cases was smaller in the event of a successful outbreak. Specifically, when 25%, 50%, and 75% of the population underwent daily screening using RT-PCR, the probability of an outbreak decreased from 0.1950 in the absence of testing to 0.1295, 0.0729, and 0.0248, respectively (**Fig. 2A**). When all individuals were screened daily, the probability of an outbreak vanished. In scenarios with the same fraction of individuals undergoing testing, the mean epidemic sizes in the event of a successful outbreak were 91.0%, 79.5%, and 50.7%, respectively, compared to 96.0% when there was no testing (**Fig. 2B**). Moreover, as the percentage of the tested population increased, we observed a wider deviation in epidemic size around its mean (**Fig. 2C**). The average extinction time, when more people participated in the screening, was slightly shortened. Conversely, in case of a successful outbreak, the time for the disease to die out was observed to be delayed by approximately a month when 75% of individuals underwent daily screening (**Fig. 2D**).

**Figure 2.**
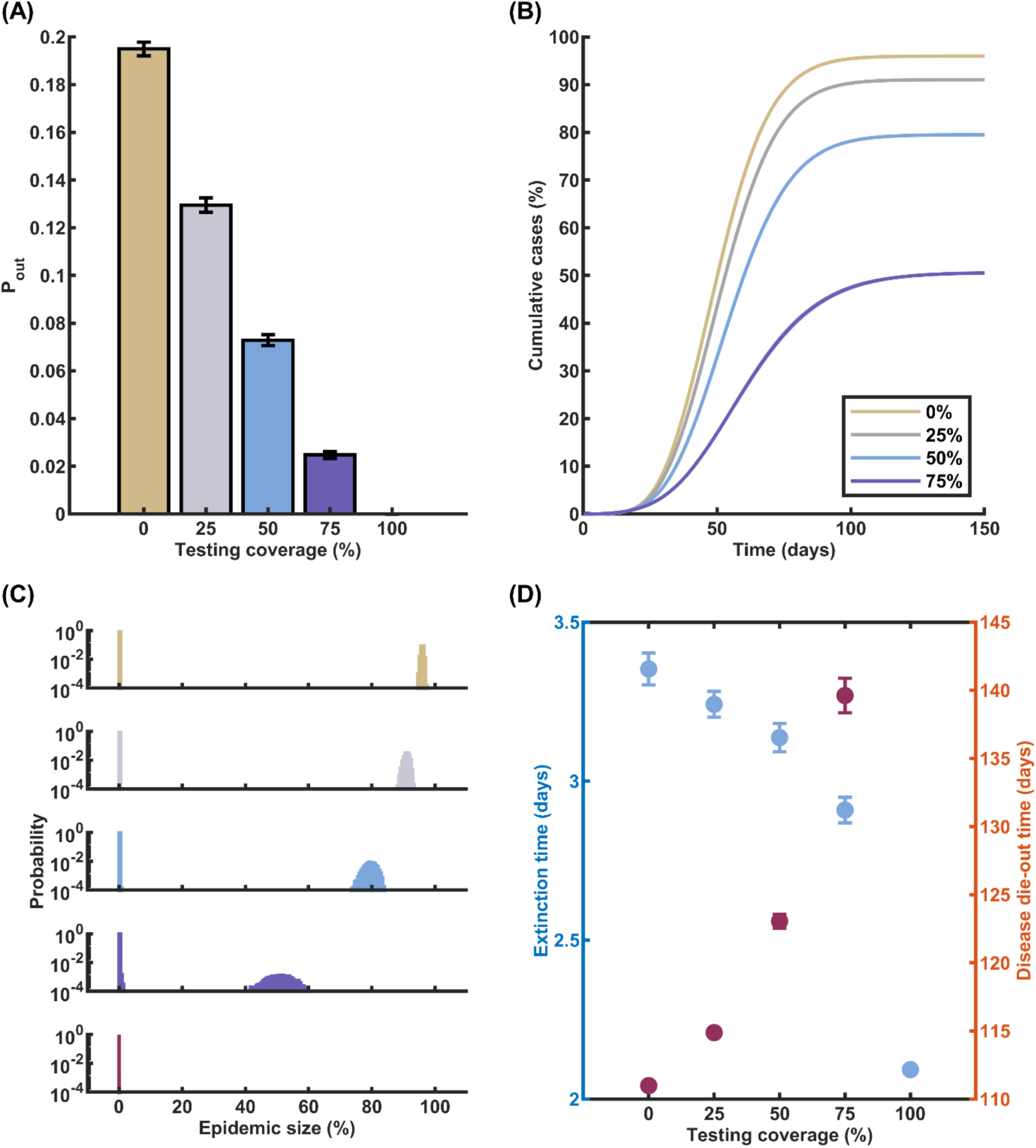
Impact of Daily Screening on Outbreak Dynamics. (**A**) Probability of an outbreak (Pout) as a function of the proportion of the population undergoing daily screening using RT-PCR. As the percentage of individuals screened daily increases, the likelihood of an outbreak decreases substantially. (**B**) Cumulative COVID-19 cases when an outbreak occurs, plotted for different proportions of the population (0%, 25%, 50%, and 75%) undergoing daily RT-PCR screening. Higher screening rates lead to a significant reduction in the total number of cases during an outbreak. (**C**) Histogram displaying the distribution of epidemic sizes across multiple simulation runs. The histogram shifts towards smaller epidemic sizes as the proportion of the population screened daily increases. (**D**) Comparison of the average time until outbreak extinction (blue circles, left axis) and the average duration of an outbreak in the event of successful disease propagation (red circles, right axis) for various daily screening rates. Increasing the proportion of the population screened daily results in faster outbreak extinction.

We further investigated the impact of the limit of detection (LOD) and turnaround time associated with the testing assays on the epidemic burden. In cases where an outbreak occurred, daily screening of 25% and 50% of the population using any assay did not result in a significant difference in epidemic size. However, these testing levels did influence the probability of an outbreak. Notably, daily screening using RT-PCR and RT-LAMP demonstrated comparable performance in terms of the probability of an outbreak. In contrast, the use of ATK on a daily basis led to a higher probability of an outbreak compared to the other assays. For instance, when 75% of the population underwent daily screening, the probabilities of an outbreak were 0.0248, 0.0246, and 0.0403 for RT-PCR, RT-LAMP, and ATK, respectively (**Fig. S1** in the Supplementary Information). These results suggest that the choice of testing assay, particularly in terms of LOD and turnaround time, can have a significant impact on the effectiveness of daily screening in mitigating the risk of COVID-19 outbreaks.

### Effectiveness of Symptom-Based Testing Strategies

While the daily screening strategy has demonstrated remarkable effectiveness in curbing disease transmission by reducing the likelihood of a successful outbreak and significantly diminishing the size of epidemics, its implementation requires a substantial volume of testing, leading to high associated costs. This financial burden may pose challenges for widespread adoption and long-term sustainability. In light of these considerations, we sought to investigate an alternative approach: a testing strategy in which tests are conducted exclusively when symptomatic individuals manifest their symptoms.

Our findings revealed that conducting tests on a larger portion of symptomatic individuals in the community can effectively decrease the probability of an outbreak and reduce the epidemic size in the event of an outbreak (**Fig. 3A** and **Fig. 3B**) Interestingly, the outcomes of the symptom-based testing strategy closely mirror those observed in the daily screening strategy. Specifically, when using RT-PCR testing on 25%, 50%, 75%, and 100% of symptomatic individuals, the probabilities of an outbreak were found to be 0.1846, 0.1555, 0.1328, and 0.1112, respectively (**Fig. 3A**), compared to the baseline scenario probability of 0.191 without any testing. Correspondingly, the epidemic sizes, when testing the same proportions of the symptomatic population, were 93.5%, 89.4%, 82.4%, and 70.0%, respectively (**Fig. 3B**). Furthermore, the results for the extinction time and the time until transmission terminated exhibited a similar trend to that observed in the daily screening strategy (**Fig. 3C** and **Fig. 3D**). These findings suggest that symptom-based testing, when conducted on a sufficient proportion of symptomatic individuals, can be an effective alternative to daily screening in terms of reducing outbreak probability and mitigating epidemic size.

**Figure 3.**
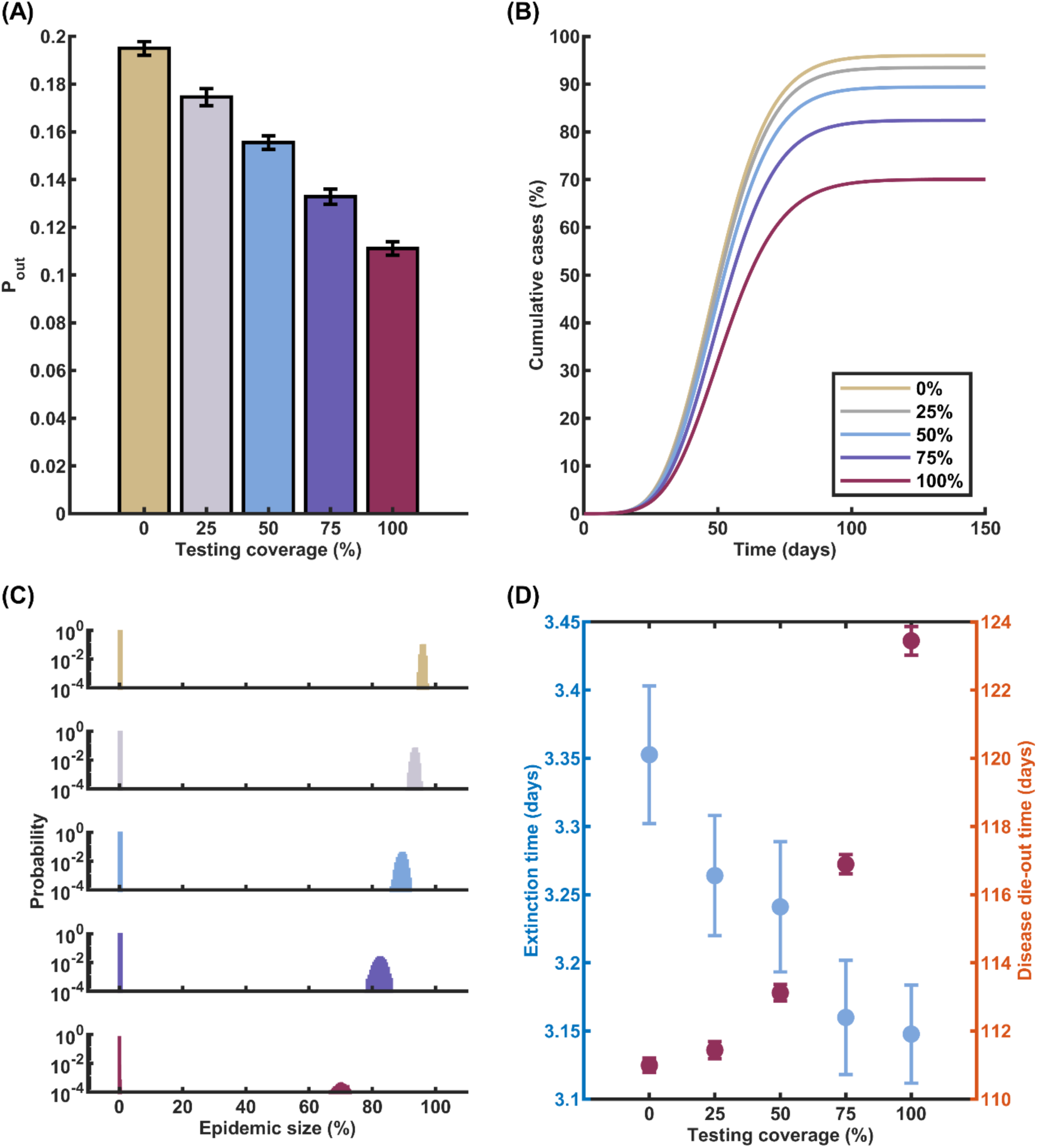
Effects of Symptom-Based Testing on COVID-19 Transmission Dynamics. (**A**) Probability of an outbreak as a function of the percentage of symptomatic individuals undergoing RT-PCR testing. (**B**) Cumulative cases in the event of a successful outbreak, with varying RT-PCR testing percentages: 0%, 25%, 50%, 75%, and 100%. (**C**) Histogram illustrating the distribution of epidemic sizes across multiple simulation runs. (**D**) Comparison of the average time until outbreak extinction (blue circles, left axis) and the average duration of an outbreak in the case of successful disease propagation (red circles, right axis) for various symptom-based testing rates.

To further evaluate the impact of different testing assays on epidemic control, we conducted additional simulations with varying LOD and turnaround times associated with RT-PCR, RT-LAMP, and ATK. Our analysis focused on a scenario in which 75% of symptomatic infectious individuals were tested. The results demonstrated that the choice of testing assay had a notable influence on the probability of an outbreak. When using RT-PCR, RT-LAMP, and ATK assays, the probabilities of an outbreak were found to be 0.1328, 0.1097, and 0.1161, respectively. This suggests that RT-LAMP provides the lowest probability of an outbreak among the three assays, followed closely by ATK and then RT-PCR. Similarly, the corresponding epidemic sizes were 82.4%, 79.3%, and 80.0% for RT-PCR, RT-LAMP, and ATK, respectively (**Fig. S2** in the Supplementary Information).

### Impacts of Integrating Contact-Based Testing with Symptom-Based Strategies

Contact-based testing has emerged as another widely adopted approach during the COVID-19 epidemic. This strategy involves identifying and testing individuals who have been in close contact with confirmed infected cases. In our research, we investigated the impact of incorporating contact-based testing as a supplementary measure alongside the primary symptom-based testing approach. Specifically, we examined a hybrid strategy where the main testing method was symptom-based, with 75% of symptomatic infectious individuals being tested using the ATK assay. Upon obtaining a positive test result from a symptomatic individual, the strategy triggered a contact tracing process within the contact network. All close contacts of the confirmed case were then subjected to daily testing for seven consecutive days, commencing as soon as the primary case’s positive test result was available. By testing the contacts promptly and repeatedly over a week, this approach aimed to quickly identify and isolate any secondary infections resulting from exposure to the primary case. The seven-day testing period was chosen to cover the potential incubation period and pre-symptomatic transmission window of COVID-19.

The results of our simulations demonstrated that the incorporation of contact-based testing alongside symptom-based testing led to a substantial reduction in both the probability of an outbreak and the cumulative number of cases. In the baseline scenario, where only symptom-based testing with ATK was employed, the probability of an outbreak was 0.1161. However, when additional testing was conducted on all close contacts of confirmed cases, the outbreak probabilities decreased significantly. Specifically, the probabilities dropped to 0.0865, 0.0874, and 0.0989 when the contacts were tested using RT-PCR, RT-LAMP, and ATK assays, respectively (**Fig. 4A**). This finding highlights the effectiveness of contact tracing and testing in identifying and isolating potential secondary infections, thereby reducing the overall transmission risk. Interestingly, despite the notable reduction in outbreak probability, the cumulative number of cases in the event of a successful outbreak remained relatively consistent at around 74%, irrespective of the testing method used for close contacts (**Fig. 4B** and **Fig. 4C**). This suggests that while contact-based testing is effective in preventing outbreaks altogether, it may have a limited impact on the final size of an epidemic once it takes hold. Nevertheless, the significant decrease in outbreak probability underscores the value of integrating contact-based testing into the overall testing strategy to enhance COVID-19 containment efforts.

**Figure 4.**
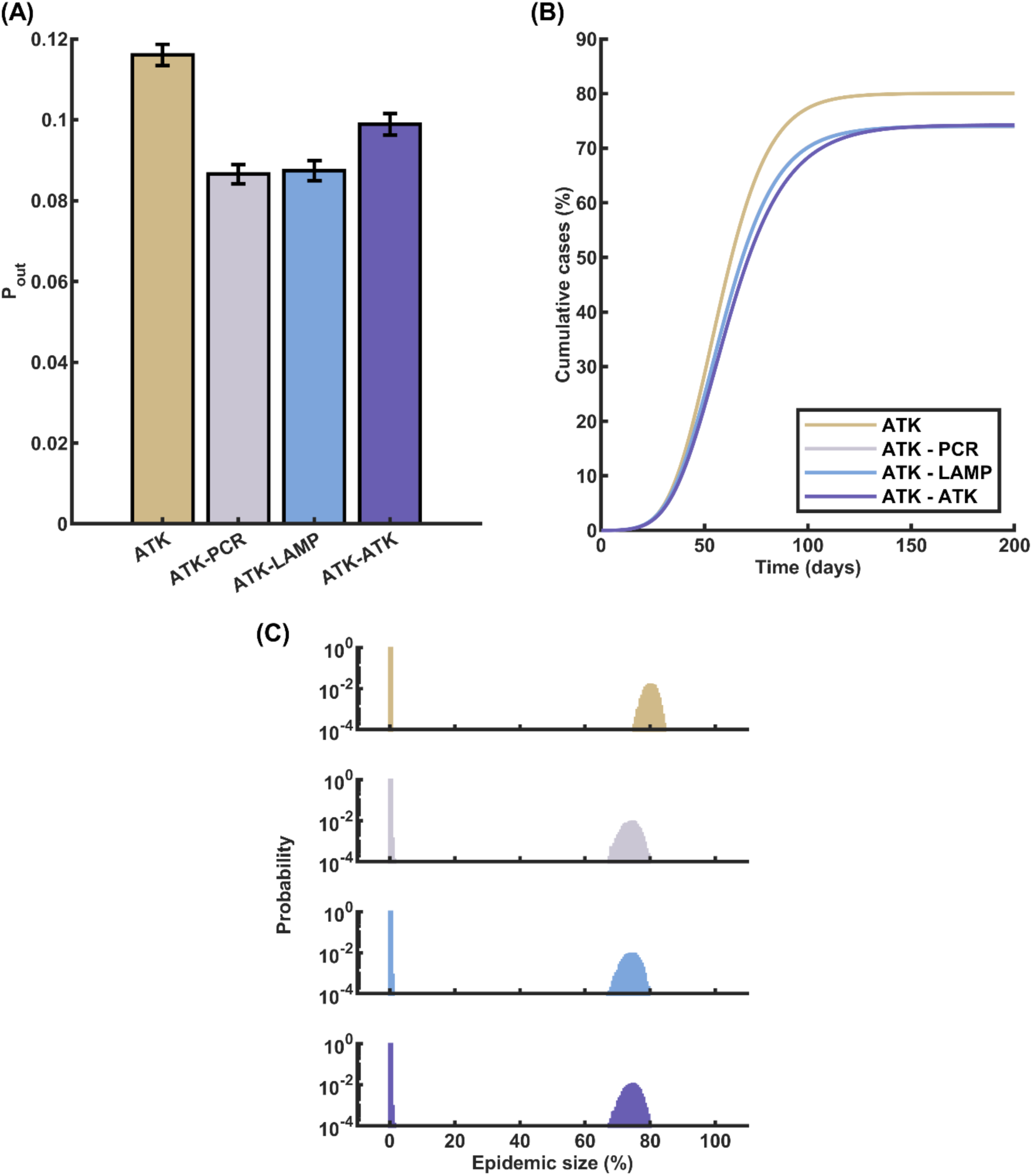
Effects of Combining Contact-Based Testing with Symptom-Based Testing on COVID-19 Outbreak Dynamics. (**A**) Probability of an outbreak when incorporating contact-based testing alongside symptom-based testing. The baseline scenario represents symptom-based testing with 75% of symptomatic individuals tested using ATK. The addition of contact-based testing using RT-PCR, RT-LAMP, or ATK for all contacts of confirmed cases results in a notable decrease in the likelihood of an outbreak compared to the baseline. (**B**) Cumulative COVID-19 case count in the event of a successful outbreak for the baseline scenario and the three contact-based testing strategies. The incorporation of contact-based testing leads to a consistent reduction in the total number of cases during an outbreak, regardless of the testing method used for contacts. (**C**) Histogram of epidemic sizes for different contact-based testing approaches. The four panels, from top to bottom, represent contact-based testing using ATK alone, ATK for symptom-based testing and RT-PCR for contact-based testing, ATK for symptom-based testing and RT-LAMP for contact-based testing, and ATK for both symptom-based and contact-based testing. The histograms demonstrate that the inclusion of contact-based testing shifts the distribution of epidemic sizes towards lower values, indicating better containment of outbreaks.

### Cost-Effectiveness Analysis of Testing Strategies for Outbreak Control

To evaluate the cost-effectiveness of each testing strategy, we conducted a comprehensive analysis of the associated expenses. Our findings revealed that, among all the strategies considered, daily screening incurred the most significant costs. This can be attributed to the repetitive nature of daily testing and the potential inefficiencies arising from its frequent implementation. However, it is crucial to acknowledge that daily screening demonstrated remarkable effectiveness in identifying infectious individuals before they exhibit symptoms or remain asymptomatic, which is a key advantage in controlling the spread of the virus.

The cost of controlling an outbreak through daily screening varied depending on the percentage of the population tested and the testing assay employed. When using RT-PCR for daily screening, the costs per individual were found to be 200, 318, 253, and 117 USD for testing 25%, 50%, 75%, and 100% of the population, respectively (**Fig. 5A**). Notably, the costs were comparatively lower when utilizing RT-LAMP and ATK testing methods for daily screening. However, it is important to consider that ATK testing carried a slightly higher risk of outbreaks compared to RT-PCR and RT-LAMP, highlighting the trade-off between cost and effectiveness.

**Figure 5.**
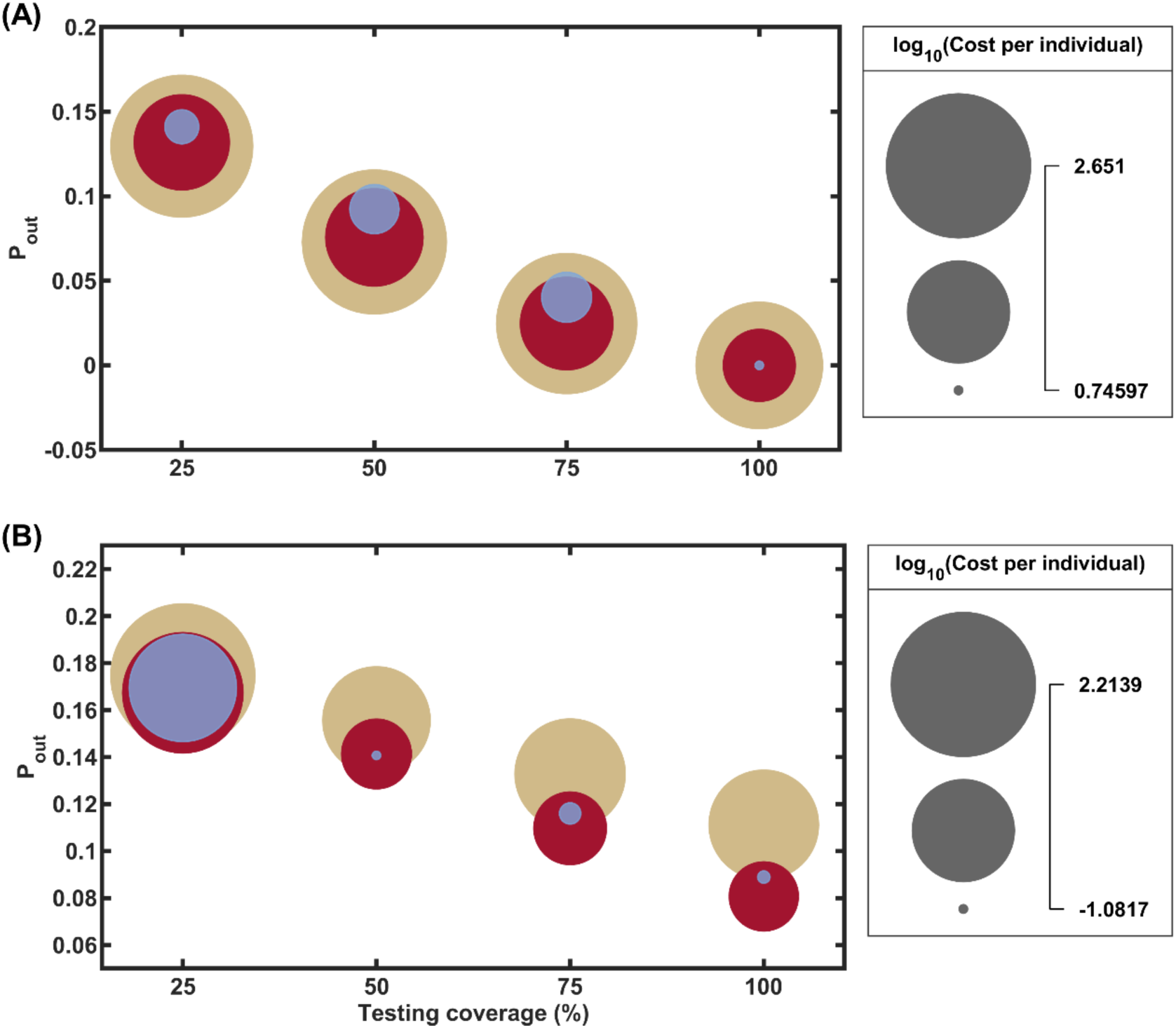
Comparison of Outbreak Probabilities and Testing Costs for Different Strategies and Assays. (**A**) Probability of an outbreak under daily screening strategies with varying percentages of the population tested (25%, 50%, 75%, and 100%) using RT-PCR, RT-LAMP, and ATK assays. The circle sizes represent the logarithmic scale of the associated testing costs per individual in USD. (**B**) Probability of an outbreak under symptom-based testing strategies with different proportions of symptomatic individuals tested (25%, 50%, 75%, and 100%) using the three assays. The marker colors indicate the type of assay used: brown for RT-PCR, red for RT-LAMP, and purple for ATK. The figure illustrates the trade-off between reducing the outbreak probability and testing costs for each strategy and assay combination.

In contrast, symptom-based testing emerged as a more cost-effective approach. When employing RT-PCR for symptom-based testing, the costs were approximately 1 USD per individual (**Fig. 5B**), while ATK testing costs were even lower, in the order of 0.01 USD per individual. These findings suggest that symptom-based testing offers a more economically viable option for outbreak control, particularly when resources are limited.

### Impact of Population Immunity on Outbreak Risk and Testing Strategies

An outbreak can exhibit varying transmission dynamics across different phases of the pandemic. For instance, in the early stage, when nearly the entire population is susceptible to the infection, a significant outbreak can be triggered by the introduction of just one primary infectious individual. However, as the outbreak progresses, the number of infected individuals increases, leading to more people becoming immunized and reducing the population’s overall susceptibility. To investigate the impact of population immunity on outbreak dynamics, we considered various initial proportions of the population in the recovered state, ranging from 30% to 80%, to reflect the levels of immunity that may be present in the later phases of the pandemic. In these simulations, we introduced a continuous inflow of infections into the system at a rate of one infectious person per day. This approach aimed to mimic the ongoing importation of cases from external sources. By incorporating this continuous introduction of new infections, we could assess the robustness of the population immunity and testing strategies in preventing new outbreaks, even in the face of persistent exposure to the virus.

In our analysis, we implemented a symptom-based testing strategy and varied the testing percentage from 25% to 100% to assess its impact on outbreak dynamics in the presence of population immunity. Our findings revealed that symptom-based testing, even with varying levels of prior population immunity, can significantly reduce the proportion of cumulative cases. In the absence of testing, the proportion of cumulative cases in the scenario with 40% initial immunity was 53.79%. However, as the percentage of symptomatic infectious individuals being tested increased, the proportion of cumulative cases decreased accordingly. Specifically, when 25%, 50%, 75%, and 100% of symptomatic infectious individuals were tested, the proportion of cumulative cases reduced to 50.43%, 45.44%, 37.85%, and 25.12%, respectively (**Fig. 6A**). In parallel, the percentage of isolated individuals increased to 7.259%, 13.02%, 16.26%, and 14.41% when 25%, 50%, 75%, and 100% of symptomatic infectious individuals were tested, respectively (**Fig. 6B**).

**Figure 6.**
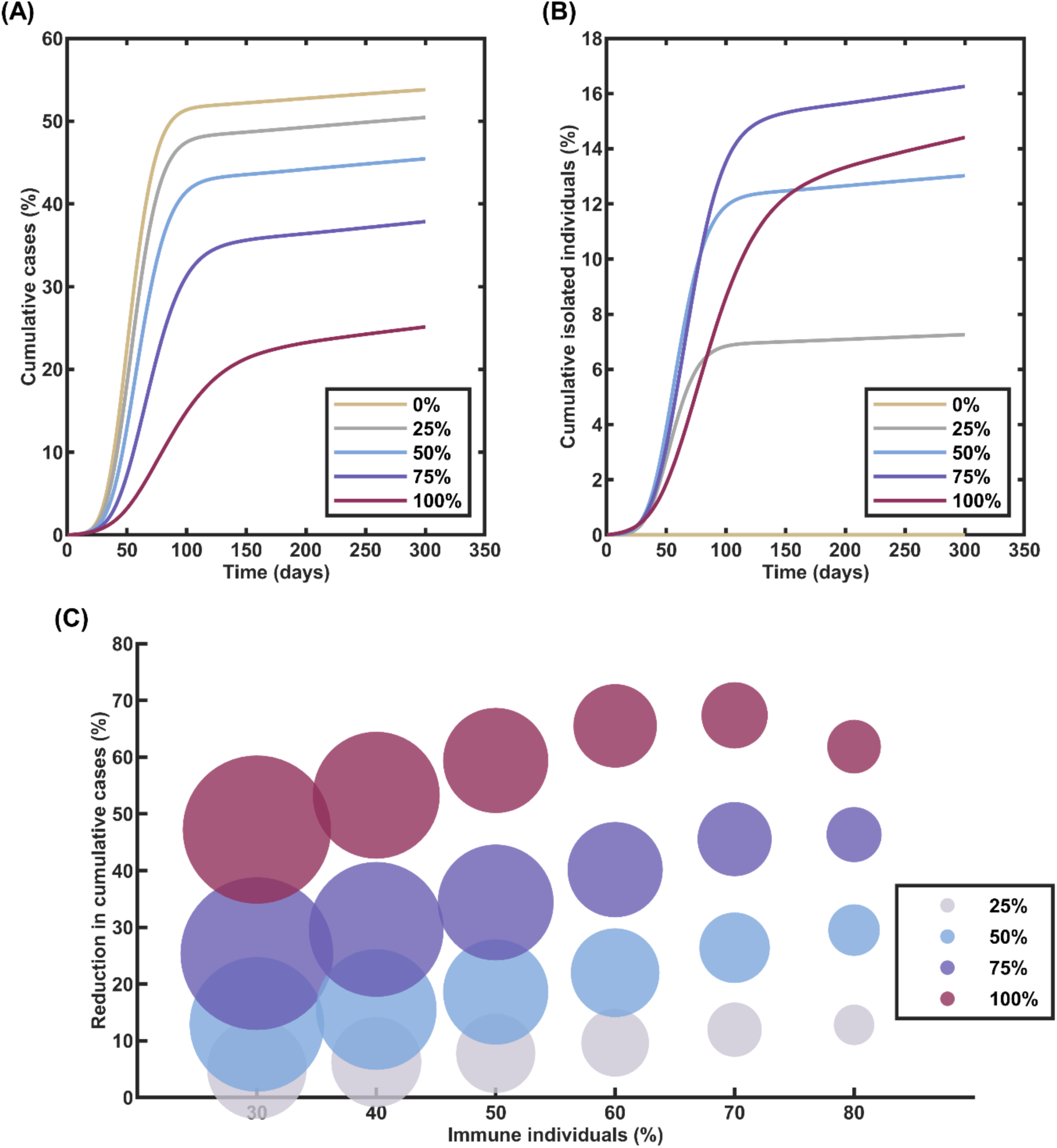
Effectiveness of Symptom-Based Testing in Scenarios with Continuous Importation of Infectious Individuals and Varying Levels of Population Immunity. (**A**) Proportion of cumulative COVID-19 cases when different percentages of symptomatic individuals (25%, 50%, 75%, and 100%) undergo testing, and 40% of the community has already acquired immunity. Increasing the percentage of symptomatic individuals tested leads to a substantial reduction in the proportion of cumulative cases, even in the presence of continuous importation of infections. (**B**) Proportion of individuals isolated under different symptom-based testing percentages when 40% of the community is immune. Higher testing rates result in a larger proportion of individuals being isolated, contributing to the containment of the outbreak. (**C**) Percentage reduction in cumulative COVID-19 cases for various proportions of symptomatic individuals undergoing testing, plotted against the proportion of the population already immune. The size of the circles represents the extent of testing utilized, with larger circles indicating higher numbers of tests employed.

Finally, our analysis revealed a relationship between the percentage of symptomatic individuals tested and the overall testing counts. Surprisingly, we found that testing 100% of symptomatic individuals required a similar amount of testing resources as testing only 50% of them. This finding challenges the intuitive understanding that higher testing coverage should necessitate a proportionally higher financial investment and that there exists an optimal level of testing coverage beyond which incremental costs may outweigh additional benefits in outbreak control. In fact, we found that when the initial immune percentage was 50%, 60%, or 70%, testing 100% of symptomatic infectious individuals required slightly lower testing quantities compared to testing 50% of them while simultaneously achieving a greater reduction in cumulative cases (**Fig. 6C**).

## Discussion

In this study, we conducted a comprehensive evaluation of various testing strategies for managing COVID-19 outbreaks, focusing on daily screening, symptom-based testing, and contact-based testing. Each of these strategies was assessed using different testing assays, including RT-PCR, RT-LAMP, and ATK, either alone or in combination. Our simulations demonstrated that daily screening was the most effective approach in mitigating the risk of a COVID-19 outbreak and reducing its overall impact once it occurred. By increasing the proportion of individuals undergoing regular testing, the likelihood of community transmission significantly decreased. However, daily screening also emerged as the most costly option among the strategies evaluated due to the high frequency of testing required. To mitigate expenses, the use of the ATK assay, which is the least expensive option, could be considered. It is important to note, however, that the ATK assay has a higher limit of detection (LOD) compared to RT-PCR, which may lead to delayed virus detection and potentially missed cases. One way to balance the cost and effectiveness of daily screening could be to reduce the testing frequency to intervals such as every 3, 5, or 7 days ^56^. This approach could help manage costs while still maintaining a reasonable level of outbreak control. Furthermore, our analysis suggests that RT-PCR and RT-LAMP assays outperform ATK in terms of reducing outbreak risk and infection rates. This advantage is likely due to their lower LOD and faster virus detection capabilities, enabling earlier identification and isolation of infected individuals.

Symptom-based testing is a commonly adopted strategy ^57,58^, owing to its lower cost compared to daily screening, despite its reduced effectiveness in mitigating outbreak risks and limiting epidemic size. Our analysis reveals that even when 100% of symptomatic infectious individuals are tested, the lowest achievable probability of an outbreak saturates at approximately 0.13. This limitation highlights the inherent challenges of relying solely on symptom-based testing, as it fails to identify asymptomatic and pre-symptomatic cases, which can contribute significantly to disease transmission ^8,59,60^. However, the cost-effectiveness of symptom-based testing makes it an attractive option, particularly in resource-constrained settings. Interestingly, our study suggests that the LOD of the testing assay is not the primary determinant of its efficacy in mitigating the disease burden. Instead, the turnaround time emerges as a crucial factor in reducing outbreak risk and infection rate. This finding emphasizes the importance of rapid testing and timely isolation of infected individuals, as delays in obtaining test results can lead to further transmission events.

Our modeling results also demonstrated that implementing additional testing on the contacts of primary cases can significantly reduce the risk of an outbreak and the overall epidemic size. By identifying and isolating infected individuals within the contact network, the chain of transmission can be effectively interrupted, limiting the spread of the virus. Interestingly, while the epidemic size remained similar across different assays when a successful outbreak occurred, both RT-PCR and RT-LAMP assays outperformed ATK in terms of reducing the overall risk of an outbreak. This superior performance can be attributed to the lower LOD of RT-PCR and RT-LAMP assays, which allows for earlier detection of the virus in infected individuals. The faster detection enables prompt isolation measures to be implemented, thereby minimizing the opportunity for further transmission. In contrast, the higher LOD of ATK may lead to delayed detection and, consequently, a higher chance of an infected individual spreading the virus before being identified and isolated.

In addition to assessing testing strategies in the early stages of an outbreak, we also investigated scenarios where a portion of the population has already acquired immunity to the disease. This situation is particularly relevant in the later stages of an epidemic when a significant number of individuals may have been infected and recovered or have received vaccinations. In these scenarios, we focused on the symptom-based testing strategy, which is commonly employed during the later phases of an epidemic ^61^. Our analysis revealed that the most effective and cost-efficient approach is to test all symptomatic infectious individuals, as this allows for the identification and isolation of active cases, thereby limiting further disease transmission. However, relying solely on symptom-based testing may not be sufficient to break the transmission chain completely, as it does not account for asymptomatic or pre-symptomatic individuals who can still spread the virus ^47,59^. To address this limitation, we found that implementing contact-based testing as a complementary measure can further reduce the transmission chain. By identifying and testing the contacts of confirmed cases, regardless of their symptomatic status, we can detect and isolate infected individuals who may have been missed by symptom-based testing alone. This combined approach of symptom-based and contact-based testing proves to be a potent strategy for controlling the spread of the disease, even in situations where a significant proportion of the population has already acquired immunity.

However, our study has some limitations that should be acknowledged. Firstly, we did not account for the possibility of reinfection, which may occur several months after the initial infection ^62^. Reinfection could potentially alter the dynamics of disease transmission and the effectiveness of testing strategies, particularly in the later stages of an epidemic. Secondly, our testing approach assumes a deterministic outcome based on the viral load and the assay’s limit of detection without considering the inherent variability in assay sensitivity ^25^. This simplification may overlook the potential for false-negative results, which could impact the effectiveness of testing strategies in real-world settings. Thirdly, we did not incorporate the heterogeneity of viral load at the individual level, which may influence the detectability of infections and the effectiveness of testing approaches ^63^. Finally, our study assumes a specific contact network structure based on a power-law distribution, which may not fully capture the complexity and diversity of real-world social interactions. The structure and properties of the contact network could influence disease transmission dynamics and the effectiveness of testing strategies ^64^. Future studies could explore the impact of different network structures and incorporate more realistic social mixing patterns to provide a more comprehensive understanding of the robustness of testing strategies across various contexts.

## Conclusions

In conclusion, our study provides a comprehensive assessment of various testing strategies for managing COVID-19 outbreaks, considering both their effectiveness and cost-efficiency. The findings reveal that daily screening is the most robust approach to minimizing the likelihood and impact of outbreaks, but it comes with the highest financial burden, necessitating strategic and targeted implementation. Our evaluation of RT-PCR, RT-LAMP, and ATK assays highlights that lower limits of detection and faster turnaround times significantly enhance outbreak control. However, this improved performance often comes at an increased expense. ATK and RT-LAMP present more cost-effective alternatives, albeit with a slightly reduced efficacy compared to RT-PCR. Symptom-based testing and contact-based testing emerge as economically viable options for disease management, particularly when used in combination. These strategies prove especially effective in curtailing transmission during the later stages of an epidemic when a significant portion of the population has acquired immunity.

Our findings underscore the importance of carefully selecting and combining testing strategies based on the specific context, stage of the epidemic, and available resources, striking a balance between disease control efficacy and economic considerations. By adapting testing approaches to the evolving epidemiological landscape, prioritizing rapid case identification and isolation, and considering the level of population immunity, public health authorities can optimize their response to COVID-19 and future infectious disease outbreaks. This study contributes to the development of evidence-based policies for pandemic preparedness and response, emphasizing the need for flexible and context-specific testing strategies that protect population health while managing limited resources effectively.

## Data availability

The authors confirm that the data supporting the findings of this study are available within the article and its supplementary information.

## Acknowledgments

The authors would like to acknowledge the financial support from the Center of Excellence on Medical Biotechnology (CEMB), The Science, Research and Innovation Promotion and Utilization Division, The Office of the Permanent Secretary Ministry of Higher Education, Science and Innovation, Thailand.

## Author contributions

Conceptualization, C.M.; Methodology, C.M., T.C., C.S., N.Y., and P.S.; Software, T.C., C.S., N.Y., and P.S.; Validation, C.M., T.C., C.S., N.Y., and P.S.; Formal Analysis, T.C., C.S., N.Y., and P.S.; Investigation, T.C., C.S., N.Y., and P.S.; Resources, C.M., Su.C., K.N., and So.C.; Data Curation, T.C., C.S., N.Y., and P.S.; Writing – Original Draft, T.C., and C.M. ; Writing – Review & Editing, C.M., T.C., C.S., N.Y., P.S., Su.C., K.N., and So.C.; Visualization, T.C., and C.M.; Supervision, C.M.; Project Administration, C.M.; Funding Acquisition, C.M.

