## Supplemental Information for "Comparative Analysis of RT-PCR, RT-LAMP, and Antigen Testing Strategies for Effective COVID-19 Outbreak Control: A Modeling Study"

* To whom correspondence should be addressed.

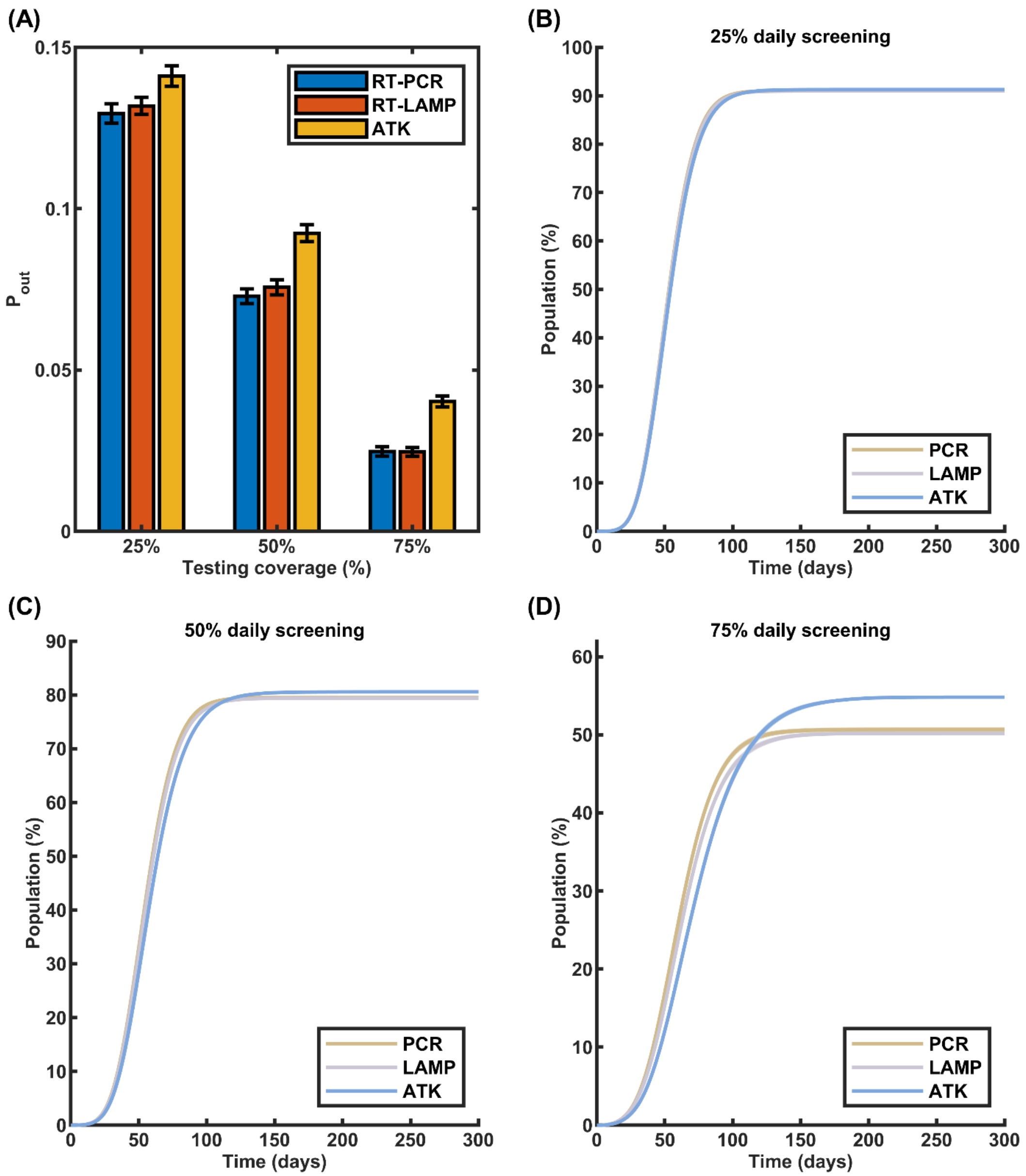


**Figure S1. Impact of limit of detection (LOD) and turnaround time on the effectiveness of daily screening strategies. (A)** Probability of an outbreak occurring under different daily screening scenarios using RT-PCR, RT-LAMP, and ATK assays. **(B) – (D)** Cumulative COVID-19 cases when an outbreak occurs, with 25%, 50%, and 75% of the population undergoing daily screening, respectively. The choice of testing assay, characterized by its LOD and turnaround time, influences the effectiveness of daily screening in reducing outbreak risk and controlling epidemic size.

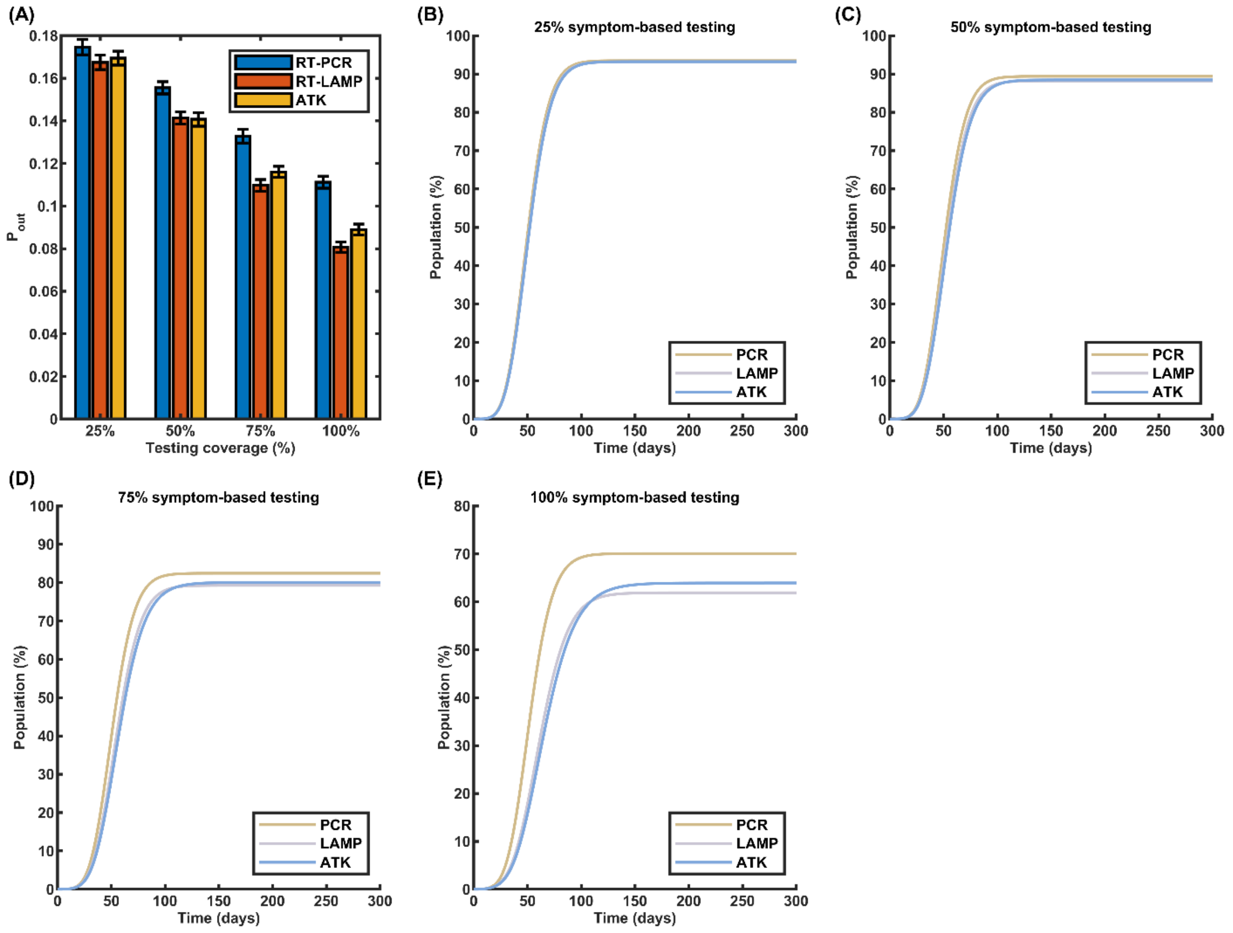


**Figure S2. Impact of limit of detection (LOD) and turnaround time on the effectiveness of symptom-based testing strategies. (A)** Probability of an outbreak occurring when different proportions of symptomatic individuals (25%, 50%, 75%, and 100%) are tested using RT-PCR, RT-LAMP, and ATK assays. **(B) – (E)** Cumulative COVID-19 cases in the event of a successful outbreak, with varying percentages of the symptomatic population undergoing testing: 25%, 50%, 75%, and 100%, respectively. The results demonstrate how the choice of testing assay, based on its LOD and turnaround time, affects the efficacy of symptom-based testing in mitigating outbreak risk and reducing the overall disease burden.
